# Cognitive decline associated with anticholinergics, benzodiazepines, and Z-drugs: findings from The Irish Longitudinal Study on Ageing (TILDA)

**DOI:** 10.1101/2020.05.09.20095661

**Authors:** Frank Moriarty, George M Savva, Carlota M Grossi, Kathleen Bennett, Chris Fox, Ian Maidment, Yoon K Loke, Nicholas Steel, Rose Anne Kenny, Kathryn Richardson

## Abstract

**Aim:** To estimate the association between patterns of anticholinergic, benzodiazepine, and Z-drug medication use and change in cognitive function in middle-aged and older adults.

**Method:** This prospective cohort study used data from the first three waves of The Irish Longitudinal Study on Ageing (TILDA), including community-dwelling adults aged ≥50 years followed for up to four years (n=7,027).

Cognitive function was assessed using the Mini Mental State Examination, animal naming test, and word recall tests. Regular medication use was self-reported at baseline and follow-up interviews at two and four years. Pharmacy dispensing claims for a subset (n=2,905) allowed assessment of medication use between interviews and cumulative dosage. Medication use at consecutive waves of TILDA was analysed in relation to change in cognitive function between waves.

**Results:** Strongly anticholinergic medications (Anticholinergic Cognitive Burden scale 3), benzodiazepines, and Z-drugs were reported by 7.3%, 5.8% and 5.1% of participants respectively at any time during the study. Adjusting for potential confounders, new anticholinergic use between interviews was associated with change in recall score (−1.09, 95% confidence interval −1.64, −0.53) over 2 years compared to non-use, but not with MMSE (0.07; 95%CI -0.21, 0.34) or animal naming (−0.70; −1.43, 0.03). The pharmacy claims analysis was consistent with this finding. Other hypothesised associations were not supported.

**Discussion:** Except for new use of anticholinergic medications, no other findings supported a risk of cognitive decline over 2-year periods in this middle-aged and older cohort. Patients and prescribers should weigh this potential risk against potential benefits of commencing anticholinergic medications.

What is already known about this subject:

- Benzodiazepines, Z-drugs and drugs with anticholinergic activity have been linked with cognitive impairment.
- Most evidence is derived from observational studies of older people, with little research on middle-aged adults.
- Medication effects may be confounded by indication and these medications are often prescribed for early symptoms of prodromal stages of dementia.

What this study adds:

- In this study of adults aged 50 years and older, no consistent pattern of change in cognitive function was associated with the medication exposures tested, except for reduced recall among new users of anticholinergic medications.
- Further research in the younger-old adult population should be conducted to confirm these findings.

## Introduction

Among the many physiological changes associated with the normal ageing process is a gradual decline in cognitive function over time.^1^ However for some individuals, this cognitive decline may be accelerated and result in mild cognitive impairment or dementia. There are no disease-modifying treatments for dementia, and so it is important to identify modifiable risk factors for cognitive decline.

Medication use is one such potential risk factor. Older people are more susceptible to adverse drug effects due to physiological changes of ageing,^2^ and often take many medications.^3,4^ The prevalence of polypharmacy in older people appears to be rapidly increasing.^5^ Drugs that target the brain or that cross the blood brain barrier and have off-target effects on the brain have received particular attention with respect to their potential impact on cognitive function.^6^

Anticholinergic medications are a diverse group of therapeutic agents, including certain antidepressants, antihistamines, drugs for urinary frequency, antipsychotics, and gastrointestinal antispasmodics. Anticholinergic effects include peripheral effects such as dry mouth and constipation, and central effects are known to include delirium and cognitive impairment.^7–12^ Cholinergic transmission is important in the brain pathways involved in cognition and memory.^13^ Older people may be particularly vulnerable to anticholinergic adverse effects on cognitive function due to age-related decreases in cholinergic neurons and receptors, and acetylcholine content of the brain.^6^ Similar evidence in relation to cognitive impairment has been presented for benzodiazepines and Z-drug hypnotics prescribed for insomnia and anxiety.^14,15^ These agents act on the gamma aminobutyric acid (GABA) A receptor, and with cholinergic transmission, GABAergic pathways are one of the most important in learning, memory, attention and executive function.^16^

Evidence in relation to long-term effects of anticholinergics and benzodiazepine and Z-drug hypnotics is primarily derived from observational studies,^9,12,14,15,17^ and relationships observed from such studies are potentially biased due to selection bias or confounding by unmeasured factors. Confounding by indication is particularly likely, as many of these medications are used for conditions that affect cognitive function, such as insomnia, anxiety, and depression. These conditions are also common in the prodromal stages of dementia.^18^ Most evidence of cognitive decline occurs in studies of the older population, where it is more likely these prescriptions were used for the early symptoms of prodromal stages of dementia. In addition, some previous studies have been cross-sectional or have included prevalent users of medications.^7^ Such approaches are unable to determine direction of any causal relationship i.e. whether the medication led to cognitive impairment, or cognitive impairment led to prescription of the medication, or whether a third factor cases both. Further, only evaluating either self-reported or prescribed/dispensed medications may lead to exposure misclassification.

Here we describe how scores on three common tests of cognitive function change over four years among a population-representative cohort of adults aged 50 and older Ireland, and how these changes are associated with new use, existing use or cessation of anticholinergic medications, benzodiazepines, and Z-drugs. By including middle-aged adults, it is less likely any associations detected are due to prescribing for symptoms of prodromal dementia. Our analysis is also strengthened by adjustment over time for conditions where these medications are indicated and being able to compare findings when medication exposures are recorded by self-report or by pharmacy dispensing claims.

## Methods

### Study design and participants

This is a cohort study and has been reported to adhere to the STrengthening the Reporting of OBservational studies in Epidemiology (STROBE) statement for cohort studies.^19^

This study included 7,027 participants from The Irish Longitudinal Study on Ageing (TILDA), a nationally representative cohort study of community-dwelling adults aged 50 years and over in Ireland.^20^ Participants are followed up every two years and at every assessment complete a computer-assisted personal interview (CAPI) with a trainer interviewer, a self-completion questionnaire, and, at waves 1 and 3, undergo a health assessment in a study centre or their own home. Detailed information on participants’ social, economic, and health circumstances are collected. Ethical approval for TILDA was provided by the Faculty of Health Sciences Ethics Committee, Trinity College Dublin.

We included TILDA participants who had at least two consecutive waves of data collected from the first three waves of TILDA from 2009 to 2014 (i.e. Waves 1 and 2, or 2 and 3). TILDA did not recruit people with severe cognitive impairments such that they could not provide informed consent, and we further excluded those who stated that they had doctor diagnosed Alzheimer’s disease, dementia, organic brain syndrome, senility, or serious memory impairment, or reported taking an anticholinesterase medication or memantine (indicating treatment for dementia).

#### Outcomes

The outcome of interest was cognitive function. Three different cognitive assessments were conducted at all waves, the Mini Mental State Examination (MMSE), an animal naming test, and an assessment of recall. The MMSE is a global measure of cognitive function which comprises 30 items (covering visuospatial, language, concentration, working memory, memory recall, and orientation cognitive domains).^21,22^ It was completed by participants during their health assessment at wave 1, and during the CAPI at waves 2 and 3. The animal naming test, included in the CAPI at all waves, is a measure of verbal fluency (which reflects language and executive function),^23,24^ in which respondents are asked to name as many different animals as they can think of in one minute. The recall test is also conducted as part of the CAPI. The interviewer reads a list of ten words, and respondents are asked to recall as many words as possible immediately (scored out of 10). The same list is then read out again and participants are asked to recall the list again (scored out of 10). Finally after several minutes and further interview questions, the respondent is asked to recall all of the words that they can (i.e. delayed recall, scored out of 10). The number of words named at both immediate recalls and the delayed recall were summed giving a total ‘recall’ score out of 30.

#### Exposures

Medications with anticholinergic effects, benzodiazepines and Z-drugs were ascertained independently from two sources, first by self-report at TILDA interviews and second through the Health Service Executive Primary Care Reimbursement Service (HSE-PCRS) pharmacy claims database.

During TILDA interviews, participants are asked to report “all medications that [they] take on a regular basis, like every day or every week”, including prescription, non-prescription, over-the-counter, herbal and alternative medications, as well as vitamins. Up to twenty medications are recorded and classified by World Health Organisation Anatomical Therapeutic Classification (WHO-ATC) code (https://www.whocc.no/atc_ddd_index/). Strength and duration of use are not recorded during the interviews. Anticholinergic medications were defined based on the Anticholinergic Cognitive Burden (ACB) scale and included any drugs classified as having ‘definite’ anticholinergic effects according to the ACB scale (i.e. a score of 3).^25^ Complete lists of exposure medications (anticholinergics, benzodiazepines and Z-drugs) and their corresponding ATC codes are included in eTable 1.

HSE-PCRS administrative data on pharmacy claims is available for the subset of TILDA participants eligible for the General Medical Services (GMS) scheme and who consented to this data linkage. The GMS scheme is a form of public health cover that provides health services including prescribed medications with a small co-payment.^26^ Eligibility is based on household income and age, and so over represents older age groups and those with low incomes. Although only available for a subset of 2,905 participants, HSE-PCRS data includes details on the exact date of prescription, quantity and dose of all prescribed medication, with records prior to TILDA recruitment. This enables quantitative assessments of exposure based on the WHO Defined Daily Dosages (DDDs).

#### Covariates

We considered other characteristics which could confound the relationship between medication exposure and cognitive decline; these are fully described in eTable 2. These included demographics, self-reported incontinence, pain, and sleep difficulties, reported doctor-diagnosed eye, cardiovascular and other health conditions, as well as self-rated vision and hearing, screened depressive symptoms, and smoking status.

### Statistical methods

First, descriptive statistics were generated for baseline characteristics (i.e. measured at wave 1) for all included participants, and separately for those who had any anticholinergic, benzodiazepine, or Z-drug use during the study.

Analysis was conducted on the basis of 2-year exposure period i.e. from wave 1-2, and from wave 2-3. We calculated the 2-year change in the cognitive outcome measure over this period, and summarised this using mean and standard deviation by pattern of medication exposure. For each medication class, we considered four categories of exposure: none (no use at either wave), new (no use at baseline wave and use at follow-up wave), discontinued (use at baseline wave and no use at follow-up wave), and recurrent (use at both waves).

Assuming initiating the medications has an immediate negative impact on cognitive function, we hypothesised that (a) new use would result in greater cognitive decline relative to non-use and that (b) discontinued use would result in improved cognitive function relative to non-use. If prolonged use led to a further decline then we would observe (c) recurrent users having a greater cognitive decline compared to non-users. So to test these hypotheses each exposure group (new, discontinuing, recurrent) was compared to the non-user group.

As well as describing average change we estimated a linear regression model for change in each cognitive measure. All exposure variables were entered simultaneously. First these were adjusted for baseline wave (i.e. whether the period was wave 1-2 or 2-3), age at baseline wave, and sex, and then were adjusted for these and all other covariates listed in eTable 2. All covariates at baseline were adjusted for, as well as covariates at follow-up for potential indications for study drug classes (sleep difficulties, anxiety, depression, manic depression, other psychiatric conditions, incontinence, Parkinson’s disease). Models were estimated with robust standard errors accounting for clustering of repeated exposure periods within participants.

As a secondary analysis we repeated the main analyses but with ‘ACB3’ exposure variables (new, discontinuing and recurring) sub-divided into antidepressant, urological, antipsychotic, and ‘other’ anticholinergic classes, adjusted for covariates and patterns of benzodiazepine and Z-drug use. We tested for equality of class effects i.e. whether the parameter estimate for new use was equal across antidepressants, urologicals, antipsychotics, and other anticholinergics. This was performed using the ‘testparm’ function in Stata using the ‘equal’ option.

We used a similar analytical approach using medication exposure based on pharmacy claims data. Dispensing was captured in the 90 days before individuals’ CAPI at each wave, as well as in the period between data collection and 90 days prior to the next wave. Exposure was classified as either non-use, new, discontinued, recurrent, or temporary use, as illustrated in eFigure 1. We also considered cumulative use among recurrent users based on the number of DDDs dispensed between baseline and follow-up, divided by quartiles into four groups, to investigate possible dose-response relationships.

We used Stata (version 14.0) for analysis. To account for the multiple outcomes tested, for final adjusted effect estimates we used the Benjamini-Hochberg procedure to control the false discovery rate (i.e. the proportion of rejected null hypotheses that are incorrect rejections) to less than 5% ^27^

#### Missing data

An additional missing category was created for categorical covariates where the proportion of missing values was greater than 0.1%. For covariates with lower proportions of missing values, we used the last value carried forward, or if unavailable, first value carried back.

## Results

This study included 7,027 participants from TILDA, who contributed 13,161 2-year exposure periods. Selected descriptive characteristics are presented in Table 1 for all participants (with full details in eTable 3). Overall, the median age at baseline was 61 years (interquartile range 55 to 69 years) and the majority (55.6%) were female. The most prevalent self-reported doctor-diagnosed conditions were high cholesterol (38.3%), high blood pressure (35.5%) and arthritis (26.4%).

**Table 1.**
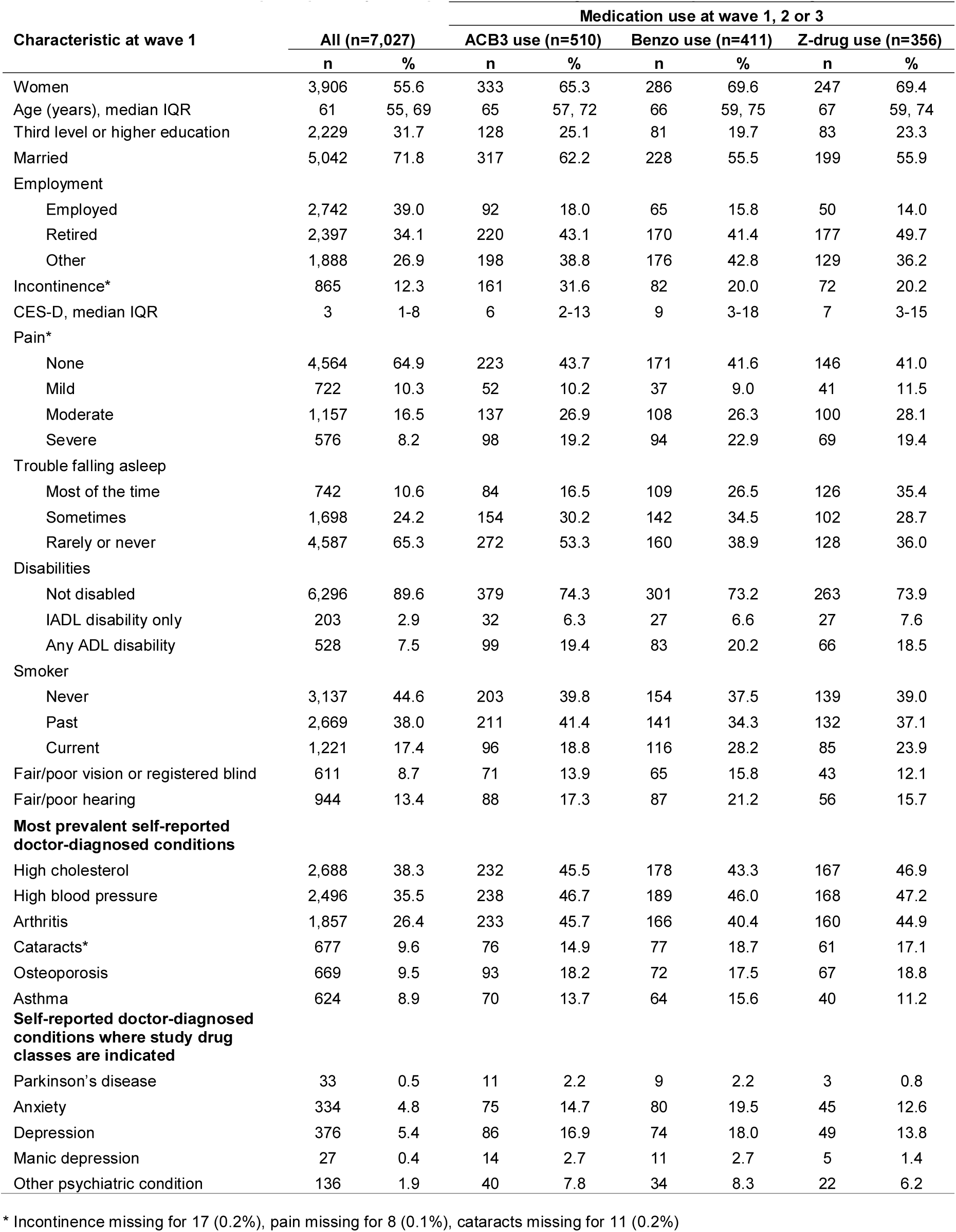
Characteristics of TILDA participants by self-reported anticholinergic, benzodiazepine, and Z-drug use.

### Self-reported medication exposure

Anticholinergic use was reported at any time during the study by 510 (7.3%) participants, whilst benzodiazepine and Z-drug use was reported by 411 (5.8%) and 356 (5.1%) participants, respectively. Those reporting these medications were more often women (65-70% compared to 56% in the complete sample) and were around 5 years older (Table 1). Users reported a substantially higher prevalence of the conditions for which medications were indicated (e.g. sleep trouble more prevalent among those reporting benzodiazepines and Z-drugs, urinary incontinence and depression in those reporting strong anticholinergic use). Those reporting anticholinergic, benzodiazepine or Z-drug use were more likely to be disabled (20% reporting an activity of daily living disability compared to 7.5% in the complete sample).

Regarding patterns of use, 267 (2.0%) participants reported new anticholinergic use at wave 2 or 3, 184 (1.4%) reported discontinued use at wave 2 or 3, and 356 (2.7%) reported recurrent use between either waves 1 and 2 or waves 2 and 3. Similarly 204 (1.6%) reported new benzodiazepine use, 184 (1.4%) discontinued use, and 248 (1.9%) recurrent use. A total of 213 (1.6%) participants reported new Z-drug use, 133 (1.0%) reported discontinued use, and 200 (1.5%) recurrent use.

The mean (standard deviation) change in cognitive measure scores across each two-year period was 0.06 (1.76) for MMSE, -1.19 (5.98) for animal naming, and -0.04 (4.40) for recall.

#### Anticholinergics

In short, no consistent pattern of decline or recovery in cognitive function was associated with any of the medication exposures we tested. In the regression models adjusted for all available patient characteristics (Table 2), new anticholinergic use was associated with a statistically significant additional decline in recall compared to no use (adjusted b coefficient -1.09, 95% confidence interval -1.64 to -0.53). Although we hypothesised that recurrent anticholinergic exposure would also be associated with reduced cognitive function, there was a relatively small decline which was not statistically significant (adjusted b coefficient -0.41, 95% CI -0.86 to 0.03). Similarly, new use was not significantly associated with additional decline in MMSE (0.07, -0.21 to 0.34) or animal naming (−0.70, −1.43 to 0.03) compared to no use, nor were recurrent or discontinued use associated with incremental changes in cognitive function.

**Table 2.** Cognitive change at 2-year follow-up compared to baseline and self-reported anticholinergic, benzodiazepine, and Z-drug medication use in TILDA.

| Medication usage | N | Baseline cognitive score mean (SD) | 2-year cognitive change, mean (SD) | b (95% CI), age, sex, wave adjusted† | b (95% CI), fully adjusted†# |
| --- | --- | --- | --- | --- | --- |
| *Anticholinergic* |  |  |  |  |  |
| **MMSE** |  |  |  |  |  |
| None | 10,863 | 28.65 (1.85) | 0.06 (1.75) | 0 | 0 |
| New | 221 | 27.90 (2.70) | 0.07 (2.11) | 0.02 (-0.25, 0.30) | 0.07 (-0.21, 0.34) |
| Discontinued | 165 | 27.82 (2.72) | 0.12 (2.00) | 0.11 (-0.19, 0.40) | 0.15 (-0.15, 0.45) |
| Recurrent | 312 | 28.02 (2.24) | -0.03 (1.84) | -0.04 (-0.22, 0.13) | 0.00 (-0.18, 0.19) |
| **Animal Naming** |  |  |  |  |  |
| None | 12,260 | 20.53 (6.60) | -1.18 (6.01) | 0 | 0 |
| New | 265 | 19.37 (7.03) | -1.91 (5.96) | -0.68 (-1.40, 0.05) | -0.70 (-1.43, 0.03) |
| Discontinued | 181 | 18.60 (6.75) | -0.60 (5.61) | 0.52 (-0.31, 1.34) | 0.47 (-0.37, 1.31) |
| Recurrent | 349 | 19.07 (6.40) | -1.23 (5.25) | -0.09 (-0.58, 0.39) | -0.13 (-0.63, 0.36) |
| **Recall** |  |  |  |  |  |
| None | 12,202 | 20.04 (5.02) | 0.00 (4.37) | 0 | 0 |
| New | 259 | 18.65 (5.59) | -1.14 (4.54) | -1.09 (-1.64, -0.53) | -1.09 (-1.64, -0.53)* |
| Discontinued | 177 | 18.57 (5.50) | -0.47 (4.78) | -0.42 (-1.13, 0.29) | -0.47 (-1.19, 0.25) |
| Recurrent | 347 | 17.94 (5.36) | -0.49 (4.82) | -0.42 (-0.86, 0.01) | -0.41 (-0.86, 0.03) |
| *Benzodiazepine* |  |  |  |  |  |
| **MMSE** |  |  |  |  |  |
| None | 11,021 | 28.64 (1.87) | 0.05 (1.74) | 0 | 0 |
| New | 176 | 27.88 (2.20) | 0.16 (2.11) | 0.12 (-0.20, 0.43) | 0.11 (-0.20, 0.42) |
| Discontinued | 155 | 27.70 (2.73) | 0.28 (2.24) | 0.27 (-0.08, 0.62) | 0.27 (-0.07, 0.62) |
| Recurrent | 209 | 27.78 (2.38) | -0.06 (1.96) | -0.07 (-0.32, 0.17) | -0.02 (-0.27, 0.22) |
| **Animal Naming** |  |  |  |  |  |
| None | 12,433 | 20.56 (6.63) | -1.19 (6.01) | 0 | 0 |
| New | 202 | 17.66 (5.97) | -0.34 (5.48) | 1.02 (0.26, 1.79) | 0.90 (0.12, 1.67) |
| Discontinued | 176 | 18.64 (6.17) | -1.55 (5.28) | -0.26 (-1.05, 0.54) | -0.43 (-1.25, 0.38) |
| Recurrent | 244 | 18.07 (5.60) | -1.32 (5.18) | -0.02 (-0.61, 0.56) | -0.09 (-0.70, 0.51) |
| **Recall** |  |  |  |  |  |
| None | 12,368 | 20.04 (5.04) | -0.04 (4.38) | 0 | 0 |
| New | 198 | 18.21 (5.18) | -0.32 (5.04) | -0.12 (-0.83, 0.58) | -0.11 (-0.81, 0.59) |
| Discontinued | 177 | 18.02 (5.62) | 0.02 (4.31) | 0.26 (-0.37, 0.88) | 0.20 (-0.43, 0.83) |
| Recurrent | 242 | 17.52 (5.14) | 0.07 (4.97) | 0.36 (-0.19, 0.91) | 0.40 (-0.16, 0.97) |
| *Z-drug* |  |  |  |  |  |
| **MMSE** |  |  |  |  |  |
| None | 11,085 | 28.64 (1.87) | 0.06 (1.74) | 0 | 0 |
| New | 178 | 27.83 (2.52) | 0.18 (2.28) | 0.13 (-0.20, 0.46) | 0.13 (-0.19, 0.46) |
| Discontinued | 120 | 27.97 (2.37) | -0.44 (2.25) | -0.48 (-0.88, -0.08) | -0.41 (-0.79, -0.03) |
| Recurrent | 178 | 27.66 (2.42) | -0.11 (1.90) | -0.13 (-0.41, 0.16) | -0.10 (-0.38, 0.18) |
| **Animal Naming** |  |  |  |  |  |
| None | 12,517 | 20.56 (6.61) | -1.18 (6.01) | 0 | 0 |
| New | 209 | 18.00 (6.28) | -1.67 (5.57) | -0.41 (-1.16, 0.35) | -0.54 (-1.29, 0.21) |
| Discontinued | 131 | 17.81 (6.30) | -1.26 (5.10) | -0.11 (-0.99, 0.77) | -0.18 (-1.06, 0.71) |
| Recurrent | 198 | 17.39 (5.43) | -0.85 (5.50) | 0.37 (-0.30, 1.04) | 0.26 (-0.41, 0.94) |
| **Recall** |  |  |  |  |  |
| None | 12,456 | 20.03 (5.05) | -0.03 (4.39) | 0 | 0 |
| New | 203 | 17.54 (5.22) | 0.22 (4.52) | 0.48 (-0.15, 1.11) | 0.52 (-0.12, 1.15) |
| Discontinued | 128 | 18.24 (5.07) | -0.30 (4.10) | -0.06 (-0.77, 0.66) | 0.01 (-0.71, 0.72) |
| Recurrent | 198 | 17.60 (5.05) | -0.53 (4.74) | -0.22 (-0.81, 0.38) | -0.22 (-0.82, 0.38) |

† b coefficients shown are change relative to 'None' category.

Cognitive decline did not differ significantly by anticholinergic drug class (eTable 4). However, new use of an antipsychotic (adjusted b coefficient -1.44, 95% CI -2.63 to -0.25) or a urological (−0.98, −1.87 to −0.09) was associated with the greatest magnitude of additional decline in recall relative to no use.

#### Benzodiazepines/Z-drugs

There was no evidence of a decline in cognitive function for recurrent use of benzodiazepines; the adjusted b coefficients (95% CIs) for MMSE, animal naming, and recall were -0.02 (−0.27 to 0.22); - 0.09 (−0.70 to 0.51); and 0.40 (−0.16 to 0.97), respectively (Table 2). Similarly, associations for new and discontinued use with cognition did not reach statistical significance. No patterns of Z-drug use had a statistically significant association with change in cognition after adjustment for confounders and multiple testing. For example, the adjusted b coefficients and 95% CIs for recurrent use were - 0.10 (−0.38 to 0.18); 0.26 (−0.41 to 0.94); and -0.22 (−0.82 to 0.38) for MMSE, animal naming, and recall, respectively (Table 2).

### Pharmacy claims based exposure

Characteristics of the 2,905 (41.3%) participants with linked pharmacy claims data are presented in eTable 5. Compared to the full TILDA sample, these participants tended to be older (mean age 67.5 years), have lower levels of educational attainment, and higher prevalence of both self-reported and doctor-diagnosed health issues.

Across each medication class, the majority (≥99.5%) of those with no record of being dispensed the medication class also did not report use. For those with no reported use, between 6.2% (anticholinergics) and 9.7% (benzodiazepines) had new, discontinued or recurrent use based on the dispensing data. Among those with new or recurrent dispensing who reported no use, the proportion who had regular use leading up to follow-up (dispensed in the 30 days before follow-up, with at least two further dispensings in the 6 months before follow-up) ranged from 44% (anticholinergics and benzodiazepines) to 52% (Z-drugs).

#### Anticholinergics

Similar to the main TILDA analysis, new anticholinergic use was associated with a change of -1.21 (95% CI -1.86 to -0.56) in cognitive recall relative to no use (Table 3), and in addition, recurrent use was associated with a significant decline of smaller magnitude (−0.82, 95% CI -1.30 to −0.34). New use showed no significant associations with other outcomes (adjusted b coefficient 0.02, 95% CI -0.32 to 0. 36 for MMSE, and -0.25, -1.02 to 0.52 for animal naming) nor did recurrent use (MMSE 0.17, −0.08 to 0.42; animal naming -0.48, −1.04 to 0.07). Discontinued use was not significantly associated with any of the three outcomes. When categorising recurrent anticholinergic use into quartiles of cumulative DDDs dispensed between baseline and follow-up (eTable 6), there was no statistically significant linear trend across cumulative anticholinergic dose in relation to change in cognitive function.

**Table 3.** Cognitive change at 2-year follow-up compared to baseline and pharmacy claims based anticholinergic, benzodiazepine, and Z-drug medication use in TILDA.

| Medication usage | N | 2-year cognitive change, mean (SD) | b (95% CI), age, sex, wave adjusted† | b (95% CI), fully adjusted†# |
| --- | --- | --- | --- | --- |
| *Anticholinergic* |  |  |  |  |
| **MMSE** |  |  |  |  |
| None | 3,240 | 0.02 (1.99) | 0 | 0 |
| New | 180 | 0.02 (2.31) | -0.01 (-0.35, 0.34) | 0.02 (-0.32, 0.36) |
| Discontinued | 146 | 0.12 (2.03) | 0.14 (-0.18, 0.47) | 0.14 (-0.19, 0.47) |
| Recurrent | 286 | 0.15 (2.17) | 0.11 (-0.13, 0.35) | 0.17 (-0.08, 0.42) |
| Temporary | 337 | 0.08 (1.95) | 0.04 (-0.17, 0.26) | 0.05 (-0.17, 0.28) |
| **Animal Naming** |  |  |  |  |
| None | 3,769 | -1.09 (5.75) | 0 | 0 |
| New | 213 | -1.43 (5.42) | -0.30 (-1.06, 0.46) | -0.25 (-1.02, 0.52) |
| Discontinued | 163 | -1.14 (5.18) | -0.13 (-0.94, 0.68) | -0.25 (-1.07, 0.57) |
| Recurrent | 335 | -1.43 (5.17) | -0.39 (-0.94, 0.16) | -0.48 (-1.04, 0.07) |
| Temporary | 390 | -1.22 (5.78) | -0.21 (-0.79, 0.36) | -0.24 (-0.84, 0.35) |
| **Recall** |  |  |  |  |
| None | 3,734 | -0.13 (4.77) | 0 | 0 |
| New | 211 | -1.36 (4.55) | -1.24 (-1.87, -0.60) | -1.21 (-1.86, -0.56)* |
| Discontinued | 161 | -0.18 (4.63) | -0.01 (-0.73, 0.71) | -0.05 (-0.79, 0.68) |
| Recurrent | 330 | -0.94 (5.01) | -0.84 (-1.31, -0.38) | -0.82 (-1.30, -0.34)* |
| Temporary | 384 | -0.32 (4.74) | -0.23 (-0.73, 0.27) | -0.22 (-0.72, 0.29) |
| *Benzodiazepine* |  |  |  |  |
| **MMSE** |  |  |  |  |
| None | 3,066 | 0.00 (1.95) | 0 | 0 |
| New | 156 | 0.18 (2.60) | 0.14 (-0.28, 0.56) | 0.18 (-0.23, 0.60) |
| Discontinued | 176 | 0.07 (2.07) | 0.09 (-0.22, 0.39) | 0.07 (-0.24, 0.38) |
| Recurrent | 359 | 0.23 (2.13) | 0.18 (-0.03, 0.38) | 0.24 (0.03, 0.45) |
| Temporary | 432 | 0.07 (2.11) | 0.02 (-0.18, 0.22) | 0.06 (-0.14, 0.26) |
| **Animal Naming** |  |  |  |  |
| None | 3,560 | -1.16 (5.72) | 0 | 0 |
| New | 171 | -1.58 (5.48) | -0.40 (-1.26, 0.46) | -0.39 (-1.25, 0.47) |
| Discontinued | 205 | -0.85 (5.98) | 0.31 (-0.55, 1.16) | 0.11 (-0.76, 0.97) |
| Recurrent | 434 | -1.17 (5.13) | 0.15 (-0.33, 0.62) | 0.08 (-0.43, 0.58) |
| Temporary | 500 | -0.96 (5.80) | 0.24 (-0.29, 0.77) | 0.17 (-0.36, 0.70) |
| **Recall** |  |  |  |  |
| None | 3,519 | -0.25 (4.77) | 0 | 0 |
| New | 170 | -0.51 (4.30) | -0.15 (-0.82, 0.51) | -0.13 (-0.81, 0.55) |
| Discontinued | 202 | -0.32 (4.54) | 0.02 (-0.62, 0.65) | -0.04 (-0.70, 0.61) |
| Recurrent | 430 | -0.33 (4.78) | 0.03 (-0.38, 0.43) | 0.03 (-0.38, 0.44) |
| Temporary | 499 | -0.13 (5.09) | 0.06 (-0.39, 0.51) | 0.08 (-0.38, 0.53) |
| *Z-drug* |  |  |  |  |
| **MMSE** |  |  |  |  |
| None | 3,385 | 0.04 (2.00) | 0 | 0 |
| New | 143 | 0.31 (2.20) | 0.24 (-0.13, 0.61) | 0.24 (-0.14, 0.61) |
| Discontinued | 98 | -0.48 (2.11) | -0.51 (-0.93, -0.09) | -0.52 (-0.93, -0.10) |
| Recurrent | 281 | -0.16 (2.13) | -0.19 (-0.42, 0.04) | -0.16 (-0.40, 0.07) |
| Temporary | 282 | 0.25 (2.01) | 0.15 (-0.09, 0.40) | 0.17 (-0.08, 0.42) |
| **Animal Naming** |  |  |  |  |
| None | 3,944 | -1.13 (5.68) | 0 | 0 |
| New | 169 | -1.09 (5.44) | 0.19 (-0.66, 1.04) | 0.17 (-0.69, 1.03) |
| Discontinued | 116 | -0.73 (5.79) | 0.45 (-0.62, 1.53) | 0.52 (-0.56, 1.60) |
| Recurrent | 325 | -1.14 (5.47) | 0.03 (-0.52, 0.57) | -0.05 (-0.60, 0.50) |
| Temporary | 316 | -1.41 (5.94) | -0.31 (-0.98, 0.37) | -0.29 (-0.97, 0.39) |
| **Recall** |  |  |  |  |
| None | 3,907 | -0.25 (4.78) | 0 | 0 |
| New | 166 | -0.79 (4.63) | -0.46 (-1.20, 0.28) | -0.55 (-1.30, 0.20) |
| Discontinued | 115 | -0.50 (5.46) | -0.11 (-1.11, 0.89) | -0.19 (-1.18, 0.80) |
| Recurrent | 319 | -0.11 (4.55) | 0.31 (-0.15, 0.77) | 0.29 (-0.18, 0.77) |
| Temporary | 313 | -0.10 (4.74) | 0.11 (-0.41, 0.63) | 0.05 (-0.48, 0.59) |

† b coefficients shown are change relative to 'None' category.

#### Benzodiazepines/Z-drugs

Again, as in the main analysis, there were no statistically significant changes in cognitive outcomes associated with patterns of benzodiazepine or Z-drug dispensings (Table 3). For example, recurrent use of benzodiazepines relative to no use had beta coefficients after adjustment of 0.24 (95% CI 0.03 to 0.45) for MMSE, 0.08 (−0.43 to 0.58) for animal naming, and 0.03 (−0.38 to 0.44) for recall.

There was a significant trend of increasing MMSE score across higher cumulative DDDs of benzodiazepines (p=0.001), where for example the highest dose quartile (greater than 736 DDDs) was associated with a 0.71 (95% CI 0.25 to 1.16) relative increase in MMSE compared to no use (eTable 6).

## Discussion

We estimated the effect of strongly anticholinergic medications, benzodiazepines and Z-drugs on three tests of cognitive function, within a large longitudinal cohort of community dwelling adults strengthened through the linkage of pharmacy claims data. Although new anticholinergic use, and to some extent recurrent anticholinergic use, were associated with statistically significant reductions in recall scores over 2-year follow-up compared to non-use, there was no evidence of significant declines in MMSE or in verbal fluency as measured by animal naming. Although the magnitude of change was small, corresponding to approximately 0.2 SDs in recall score, and may not be clinically meaningful or detectable within an individual, a change of 0.2 SD does represent a more substantial effect at the population level. There was no evidence that new or recurrent use of benzodiazepine or Z-drug use was associated with any impairment of cognitive function in this population.

### Strengths and weaknesses of the study

Our study has several strengths and limitations. We included a large sample of representative community-dwelling adults, and included participants aged 50 years and older rather than many previous studies which have only considered older age groups (typically 65 years and older).^28^ We simultaneously considered multiple medication classes for which there is evidence suggesting an effect on cognitive function and included a range of outcomes which captured multiple aspects of cognitive function. We also considered changes in exposure to medication among participants to estimate the marginal effect of commencing use or discontinuing use, providing stronger evidence of potential causality. We aimed to further strengthen the evidence for a potential causal claim by considering dose-response relationships and adjusting for likely indications for anticholinergics and hypnotics to reduce confounding by indication and protopathic bias. While we did not account for drugs with ‘possible’ anticholinergic effects (e.g. an ACB score or 1 or 2), those we did consider have established and clinically relevant anticholinergic effects, including delirium. Although anticholinergic scales differ on which medications they class as anticholinergic, they are typically consistent in classifying those with strong or definite anticholinergic activity.^29^

Our study was strengthened by using both self-reported medication use and pharmacy claims data to ascertain medication exposure, and capturing both prescribed and non-prescribed medications. Although self-reported medication use might underestimate exposure, and pharmacy claims data might over-estimate exposure, our findings were generally similar for both sources. Some individuals taking anticholinergics or hypnotics leading to cognitive decline may have been excluded from TILDA if not cognitively well at wave 1 recruitment as participants were required to provide written informed consent, potentially biasing our study towards a null finding. Although our study period of four years is relatively long compared to some studies, it may be that even longer periods of cumulative exposure are required to observe an impact on cognitive function.^30^

### Comparison with other studies

A number of previous studies have reported anticholinergic drug use associated with decline in MMSE,^10,31–33^ which was not replicated in the present study. These studies showed larger overall declines in MMSE among their study samples than observed here, for instance a 0.33 reduction associated with anticholinergic use versus non-use over two years in the Cognitive Function and Ageing Study.^31^

The relatively young population of our study, in comparison to previous research,^10,15^ may also have contributed to a lack of observed association. A recent systematic review of longitudinal studies found that among studies with high methodological quality, there was an association between anticholinergic burden and dementia or cognitive impairment, except in middle-aged adults.^28^ The lack of association in the two identified studies including adults aged 50-64 years was postulated to be due to lower dementia risk and greater baseline cholinergic brain activity than older adults. In the present study there is also a potential ceiling effect due to the high baseline MMSE scores.

A further explanation may be that the MMSE was administered in a different setting at wave 1 (at the health assessment by a trained nurse) compared to subsequent waves where it was administered during the CAPI by a trained interviewer in the participant’s home, with the potential for measurement error and selection bias. However, we adjusted for baseline wave in our analysis, and our findings were consistent when restricted to wave 2 and 3 only (results not shown).

Most previous studies on benzodiazepine and Z-drug use report declines in cognitive function, although in most cases not statistical significant.^34–38^ One of the studies with the largest magnitude of decline examined chronic benzodiazepine use (repeated use at baseline, 2-year follow-up and 4-year follow-up), suggesting a potential duration of use and/or cumulative dose effect.^36^ However, we found no evidence of greater cognitive decline with greater DDDs of benzodiazepines dispensed. Consistent with our findings, recent studies suggest no or only modest associations between recurrent benzodiazepine use and incident dementia.^15,39–42^ Non-randomised comparisons of benzodiazepine withdrawal versus continuation in studies of older adults have shown an association between withdrawal and improved cognitive function,^43,44^ but a randomised trial including adults of any age on benzodiazepines found no difference in recall six months post withdrawal.^45^

### Study implications

Our findings suggest that among a wider middle-aged and older population, there might be a shortterm impact on memory associated with new strong anticholinergic use but that this is unlikely to worsen if use is prolonged. Further research in the younger-old adult population on this issue should be conducted to confirm these findings. The lack of associations in our study could indicate lesser ability for anticholinergic medications to cross the blood brain barrier in this younger-old population or that protopathic bias is responsible for associations between anticholinergic medications and cognitive decline observed in older populations.

If our findings are confirmed, the potential risk of declines in memory should be communicated to people purchasing or being prescribed anticholinergic medicines so that, as with any medication class, the potential benefits and harms can be weighed up. For possible new users, prescribers should consider alternative therapeutic options, including non-pharmacological treatments. Although risks of recurrent use of anticholinergics were not supported by our findings, prescribers may wish to consider ongoing need and potential for deprescribing given other published evidence on the impact of anticholinergics on cognitive decline in older ages with prolonged exposure.^10,31–33^

Research on the cognitive effects of anticholinergics and hypnotics has been predominantly observational, and causal inference is complicated by the extremely close links between illness and medication exposure. Future interventional studies should consider the cognitive effects of deprescribing anticholinergics, benzodiazepines, or Z-drugs compared to continuation of these agents. Such studies would provide stronger evidence to inform clinician and patient decisions to continue these medications.

## Data Availability

The data that support the findings of this study are available from TILDA, with further details on access available at https://tilda.tcd.ie/data/accessing-data/.

## Acknowledgement

We wish to acknowledge all of the participants in The Irish Longitudinal Study on Ageing (TILDA), and the Health Service Executive Primary Care Reimbursement Service (HSE-PCRS) for providing access to the administrative pharmacy claims data used in this study.

## Conflict of interest statement

The authors declare that they have no competing interests, other than IM has received personal fees for guest lectures and to support travel from Astellas Pharmaceuticals; YL reports personal fees from Thame Pharmaceuticals, NC and CF have received grants and personal fees from Astellas Pharmaceuticals.

## Funding information

This work was supported by the UK Alzheimer’s Society [AS-PG-2013-017], Funding for TILDA is provided by the Irish Government, the Atlantic Philanthropies, and Irish Life.

## Author contributions

GS, CG, KB, CF, IM, YL, NS, RAK, and KR conceived the study, FM, GS, CG, RAK and KR designed the study, KB and RAK acquired the data, FM, GS and KR analysed and interpreted the data, FM drafted the manuscript and all authors were involved in the critical revision of this.

